# Antibiotic prescribing for lower UTI in elderly patients in primary care and risk of bloodstream infection: a cohort study using electronic health records

**DOI:** 10.1101/2020.03.11.20033811

**Authors:** Laura Shallcross, Patrick Rockenschaub, Ruth Blackburn, Irwin Nazareth, Nick Freemantle, Andrew Hayward

**Affiliations:** Institute of Health Informatics, UCL; Health Data Research UK; Department of Primary Care and Population Health, UCL; Institute of Clinical Trials and Methodology, UCL; Institute of Epidemiology & Healthcare, UCL

**Keywords:** UTI, primary care, antibiotic, antimicrobial stewardship, health policy

## Abstract

**Background:** Research has questioned the safety of delaying or withholding antibiotics for suspected urinary tract infection (UTI) in older patients. We evaluated the association between antibiotic treatment for lower UTI and risk of bloodstream infection (BSI) in adults aged ≥65 years in primary care.

**Methods:** We analysed primary care records from patients aged ≥65 years in England with community-onset UTI using the Clinical Practice Research Datalink (2007-2015) linked to Hospital Episode Statistics and census data. The primary outcome was BSI within 60 days, comparing patients treated immediately with antibiotics and those not treated immediately.

**Findings:** 147,334 patients were included representing 280,462 episodes of lower UTI. BSI occurred in 0·4% (1,025 / 244,963) of UTI episodes with immediate antibiotics versus 0·6% (228 / 35,499) of episodes without immediate antibiotics. The odds of BSI were equivalent in patients who were not treated with antibiotics immediately and those who were treated on the date of their UTI consultation (adjusted odds ratio aOR 1·13; 95%-CI: 0·97-1·31). However, delaying or withholding antibiotics was associated with increased odds of death in the subsequent 60 days (aOR 1·17; 95% CI: 1·09-1·26).

**Interpretation:** Evidence on the safety of delaying or withholding antibiotics in older adults with suspected UTI is conflicting. Given the prevalence of asymptomatic bacteriuria in this population, their risk of antibiotic-related side effects, and the public health need to tackle antibiotic resistance, we recommend a trial to address this uncertainty.

## 1 Introduction

Urinary tract infections (UTIs) are common in older adults in both primary and secondary care,^1^ with *E. coli* as the causative pathogen in 70-95% of cases.^2^ The clinical spectrum of UTI ranges from mild urinary symptoms to urosepsis, but the rate of *E. coli* bloodstream infection is highest in the oldest age groups (758.5/100,000 in ≥85 years versus 53·4/100,000 in 45-64 year olds).^3^

Identifying cases of UTI can be challenging, particularly in the elderly who often present with atypical signs and symptoms of infection.^4^ Diagnostic uncertainty is compounded by the increased prevalence of asymptomatic bacteriuria in older adults (>20% in women aged ≥ 65 years compared to 5% of younger women)^5,6^ and widespread use of urine dipstick testing across healthcare settings, despite its poor positive predictive value for bacteriuria.^7^ Older patients are also at disproportionate risk of toxicity from antibiotics, as well as complications such as *Clostridium difficile* infection,^8^ adding to the complexity of management decisions.

UTI is the second commonest reason for antibiotics to be prescribed in primary care but an estimated 40-50% of these prescriptions are inappropriate.^9^ A wide range of national initiatives aiming to tackle inappropriate prescribing have reduced total prescribing by 13·2% between 2013 and 2017,^10^ and achieved reductions in broad-spectrum prescribing, even in elderly populations.^11^ However, rates of gram-negative bloodstream infections (BSI) continue to rise10 and whilst it is anticipated that reductions in prescribing will have a beneficial impact on rates of antibiotic resistance and *C. difficile* infection, this has to be balanced against the risk of increasing rare but severe outcomes such as BSI.

The safety of delaying or withholding antibiotic treatment for suspected UTI in older adults in primary care has recently been called into question by an electronic health record study by Gharbi *et al*. This study reported a 7-8 fold increase in the odds of BSI in the 60 days following consultation if antibiotic treatment was delayed or withheld by comparison with patients who were treated immediately (i.e. on the date of their first UTI consultation).^12^ Delaying or withholding antibiotics was also associated with a statistically significant increase in 60 day mortality. Gharbi *et al*. are the first to address this important research question and their findings are therefore likely to have a significant influence on policy and clinical practice, for example by reducing GPs willingness to consider the use of potentially beneficial strategies such as delayed prescribing. However, a number of research groups have strongly questioned the validity of these findings,^13^ highlighting methodological concerns around the definition of UTI episodes and the comparability and definitions of the different antibiotic treatment groups.

GPs require robust evidence on which to base empirical prescribing decisions, and in the absence of randomised controlled trials, observational studies using large-scale electronic health records can help to address this evidence-gap. We therefore attempted to replicate the findings reported by Gharbi *et al*. by analysing the same dataset and undertaking a range of sensitivity analyses to test the robustness of our findings. We addressed the following research question: In a population aged ≥ 65 years who consult primary care for suspected lower UTI, are patients who are not treated with antibiotics immediately at increased risk of BSI in the following 60 days, compared to patients who were treated with an antibiotic on the date of their consultation?

## 2 Methods

### 2.1 Database and study population

The Clinical Practice Research Datalink (CPRD) database is a nationally representative database of primary care consultations in the UK.^14^ Data in CPRD are collected anonymously from practice management systems of 674 practices and include demographic information, medical tests, diagnoses, and prescriptions. Diagnoses are entered directly by clinicians using Read codes, the main medical coding terminology in UK primary care.^15^ A subset of consenting English patients and practices (75% of English practices, 58% of all practices) are further linked to data on hospital admissions and visits to the Emergency Department from the Hospital Episodes Statistics (HES) and census data from the Office for National Statistics (ONS).

We included all patients in the CPRD-HES-ONS linked data aged 65 years or more between April 1^st^ 2007 and March 31^st^ 2015. Data were required to fulfil basic quality standards^14^ and patients entered the cohort at the latest of: the practice’s up-to-standard date, one year of continuous registration with the practice, their 65^th^ birthday, or April 1^st^ 2007. Patients left the cohort either on their date of death or 60 days before the earliest of: the practice’s last collection date, their transfer-out date, or March 31^st^ 2015.

The study was approved by the MHRA (UK) Independent Scientific Advisory Committee [ISAC-Nr.: 17 048], under Section 251 (NHS Social Care Act 2006).

### 2.2 Definition of UTI episodes

The study population comprised patients who consulted for a new episode of lower UTI that originated in the community. UTI episodes were identified from the primary care or linked hospital record using previously published codelists^12^ (Supplementary Table 1) based on Read codes and International Classification of Diseases 10^th^ revision (ICD-10).

Each patient’s earliest observed UTI code in primary and/or secondary care was set as the start date of that patient’s first episode (Figure 1). Any further UTI codes recorded within 60 days after the episode start date were considered part of the same episode. The first UTI code after 60 days was considered as the start of the next episode, in order to distinguish new episodes (most recent UTI code > 60 days before episode start) and ongoing episodes (most recent UTI code < 60 days before episode start), Figures 1A and 1B. Gharbi *et al*. used a 30 day window to distinguish between new and ongoing UTI episodes. Only new episodes of lower UTI starting in primary care were included in the analysis.

**Figure 1:**
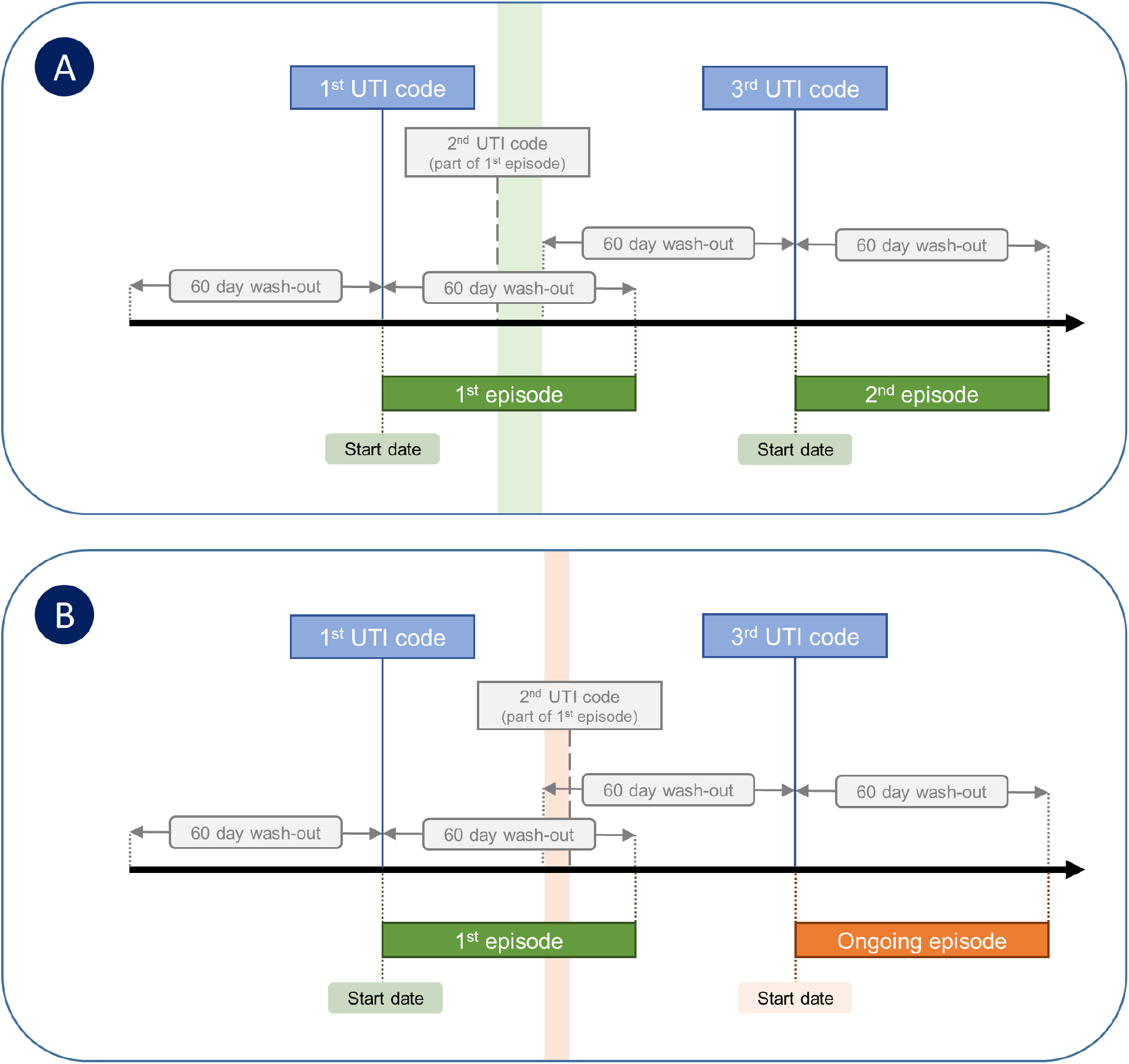
Definition of UTI episodes. The 1^st^ UTI code starts a new UTI episode (1^st^ episode). The 2^nd^ UTI code is within the wash-out period of 60 days and is considered part of the 1^st^ episode. The 3^rd^ UTI code occurs more than 60 days after the start of the 1^st^ episode and is counted as: A a new episode, because the last evidence of UTI was recorded more than 30 days before (green bar); Ban ongoing episode, because the last evidence of UTI was recorded less than 30 days before (red bar). In the latter case, the 2^nd^ UTI code falls within the 60-day wash-out period of both the 1^st^ and 3^rd^ UTI code and could have either been a consequence of the 1^st^ episode or the true start of the 2^nd^ episode. Due to this ambiguity, the 2^nd^ episode is labelled as an ongoing episode and excluded from the analysis.

We excluded episodes in which the patient was admitted to hospital, attended A&E, was referred to specialist care, or died on the day of episode start. Episodes were excluded if the linked HES record showed that the patient was in hospital on the date that the episode was recorded in primary care.

### 2.3 Exposure, outcomes and covariates

We compared patients who were immediately treated with antibiotics defined as prescription of systemic antibiotics on the same day as the episode start date, to patients who were not treated with antibiotics on the same day. In contrast to Gharbi *et al*. we considered patients who were not prescribed antibiotics and those with a delayed prescription as a single group, because delayed prescribing is not well recorded in electronic health records.

The primary outcome was BSI recorded in primary or secondary care within 60 days of the episode start date. Secondary outcomes were: all-cause mortality within 60 days; admission to hospital for reasons unrelated to UTI or sepsis within 60 days; and underlying cause of BSI. BSI was identified in primary care using Read codes and in secondary care using ICD-10 codes (which represent the primary and secondary reasons for admission) using published codelists.^12^ ICD-10 codes for sepsis were further classified as urosepsis, sepsis of other infectious origin, and unspecified sepsis (Supplementary data).

Explanatory variables included demographic characteristics: age at episode start, gender, quintile of socio-economic status (Index of Multiple Deprivation - IMD 2015), and practice region (South of England, London, East of England and Midlands, North of England and Yorkshire). We also evaluated risk factors for infection and healthcare utilisation including: Charlson comorbidity index (CCI), smoking status (non-smoker, ex-smoker, current smoker), whether the index consultation was performed as a home visit, recent hospitalisations (discharge in prior 7 and 30 days, number of admissions in prior year, total number of weeks spent in hospital in prior year), recent Accidents & Emergency attendances (attendance in prior 30 days, number of attendances in prior year), and prescription of systemic antibiotics in primary care in prior 30 days. History of recurrent UTI was defined as an explicit code for recurrent UTI, a prescription of nitrofurantoin or trimethoprim for 28 days or more (prophylactic treatment), or 2 or more consultations for UTI within a year of episode start.^12^ CCI and smoking status were calculated using all medical history in primary care before the episode start date. Patients without a smoking code were considered non-smokers. Patients whose latest record indicated a non-smoker but who had a previous record of smoking were classified as ex-smokers.

### 2.4 Statistical analysis

We undertook a univariable analysis comparing patients with and without immediate antibiotic treatment for each included variable. Continuous variables were summarised using means and standard deviations, and categorical variables using absolute numbers and proportions. Wilcoxon rank tests (continuous) and *χ*^2^ tests (categorical) were used to assess the difference between exposure groups. We tabulated diagnostic information relating to the underlying cause of sepsis for each treatment group.

Crude associations (odds ratios) between each included variable and sepsis were estimated using general estimating equations with a logit link and an exchangeable correlation structure to account for multiple UTI episodes per patient. 95%-confidence intervals (95%-CI) were calculated using Huber-White sandwich estimators. A final multivariable adjusted model was fitted including all predictors with a p-value < 0·2 in the univariable analysis. Based on earlier reviewer comments, interactions between prescribing and age or gender were also considered. The number needed to be exposed (i.e. not treated with antibiotics) to harm (NNEH) was calculated from the final model using average risk difference to adjust for covariate imbalance.^16^ 95%-CIs were estimated for the NNEH by refitting the analysis in 200 bootstrapped samples.

The same approach was used for secondary outcomes. Sensitivity analysis was undertaken restricting the follow-up/wash-out periods to 30 days, and only including the first UTI episode per patient. We also tested the sensitivity to residual confounding by performing propensity score analysis. A patient’s prior likelihood to receive treatment was estimated using multivariable logistic regression (parametric) or generalised boosted regression (non-parametric), and four different adjusted results were obtained using each set of propensity scores with either matching or inverse probability weighting.

Analysis was performed using the statistical software R version 3.6.1 for Windows.^17^ General estimating equations were fitted geepack (version 1.2-1), and propensity score analysis was performed using MatchIt (version 3.0.2) and twang (version 1.5). Code for all analyses can be found at https://github.com/prockenschaub/CPRD_UTI_sepsis_elderly.

## 3 Results

Data were available for 850,794 patients aged ≥ 65 years corresponding to 3,706,722 patient years at risk between April 1^st^ 2007 and March 31^st^ 2015 (Figure 2). The cohort included 147,334 patients with 280,462 distinct episodes of lower UTI, corresponding to 75·7 episodes per 1,000 patient-years at risk. UTI episodes mainly occurred in women 217,425/280,462 (77·5%). Most UTI episodes (244,963/280,462; 87·3%) were treated with antibiotics immediately (Table 1), and at least one antibiotic prescription was recorded in the 7 days following consultation for 6411/35499 (2·3%) UTI episodes that were not treated immediately. Factors that were associated with delayed or withheld prescribing (versus immediate treatment) included: male gender (40·9% versus 19·8%); antibiotic prescription in the previous 30 days (27·0% versus 18·2%) and GP home visits (9·6% versus 3·7%). Sepsis was recorded in 1,025/244,963 (0 4%) UTI episodes with immediate antibiotic treatment and in 228/35,499 (0·6%) episodes that were not treated immediately (Table 1). The median number of days to diagnosis of sepsis was shorter in patients who were not treated with antibiotics immediately compared to those who were treated immediately (13 days; IQR: 3-32·5 days versus 22 days; IQR: 7-40 days).

**Figure 2:**
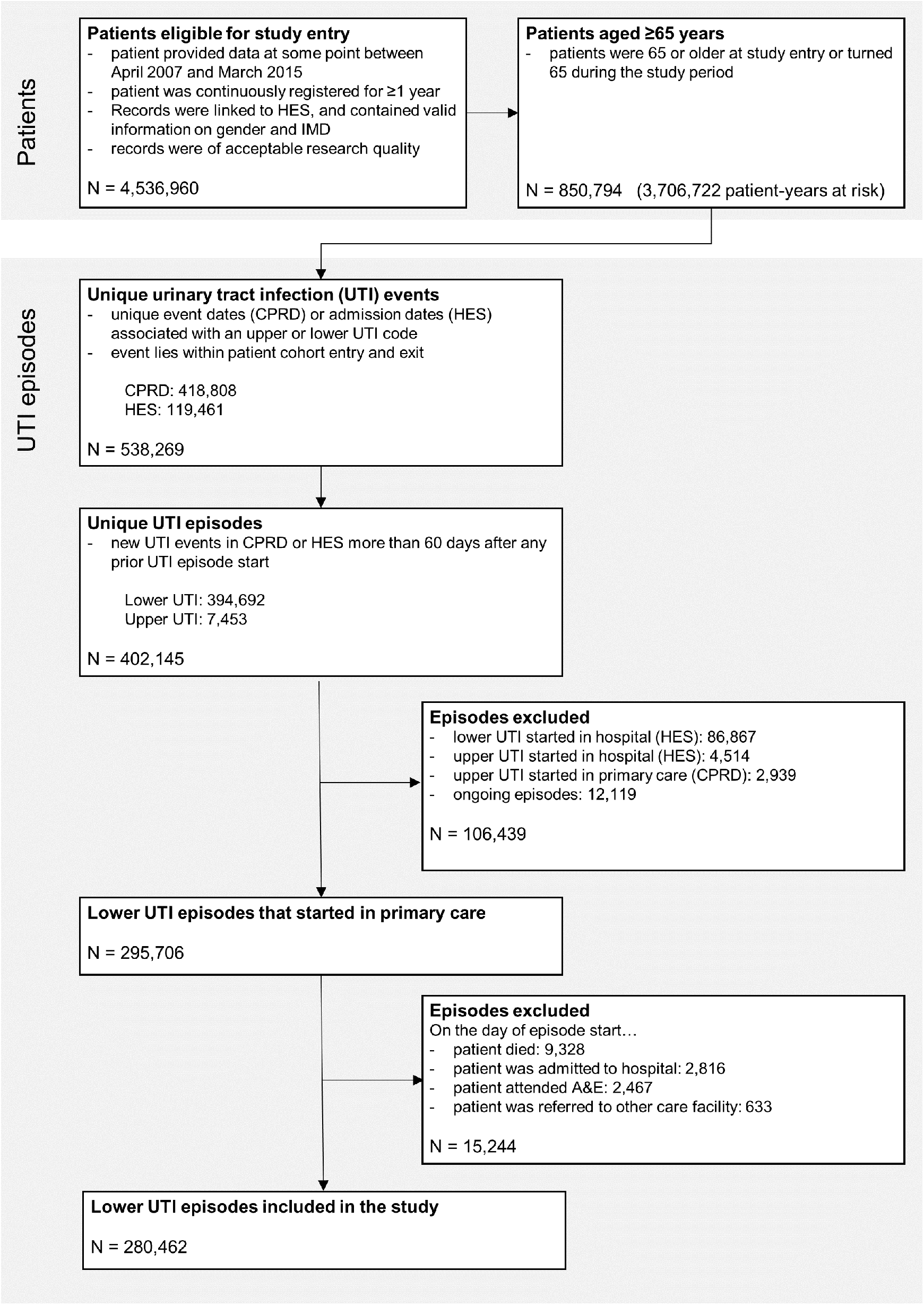
Selection of study cohort.

**Table 1:** Baseline characteristics associated with lower urinary tract infection episodes in primary care, comparing episodes with and without immediate (same day) antibiotic prescribing

| Patient characteristics | All<br>N (%) / mean (sd) | Antibiotic prescribing |  | p-value |
| --- | --- | --- | --- | --- |
|  |  | Yes<br>N (%) / mean (sd) | No<br>N (%) / mean (sd) |  |
| **Total** | 280,462 (100.0) | 244,963 (87.3) | 35,499 (12.7) |  |
| **Age (continuous)\*** | 77.94 (8.2) | 77.88 (8.1) | 78.36 (8.4) | <0.001 |
| **Age (categorical)** |  |  |  | <0.001 |
| 65-74 | 113,332 (40.4) | 99,511 (40.6) | 13,821 (38.9) |  |
| 75-84 | 106,900 (38.1) | 93,714 (38.3) | 13,186 (37.1) |  |
| ≥ 85 | 60,230 (21.5) | 51,738 (21.1) | 8,492 (23.9) |  |
| **Female** | 217,425 (77.5) | 196,459 (80.2) | 20,966 (59.1) | <0.001 |
| **Index of Multiple deprivation** |  |  |  | 0.005 |
| Q1 (least deprived) | 69,516 (24.8) | 60,482 (24.7) | 9,034 (25.4) |  |
| Q2 | 68,320 (24.4) | 59,654 (24.4) | 8,666 (24.4) |  |
| Q3 | 62,324 (22.2) | 54,607 (22.3) | 7,717 (21.7) |  |
| Q4 | 46,119 (16.4) | 40,404 (16.5) | 5,715 (16.1) |  |
| Q5 (most deprived) | 34,183 (12.2) | 29,816 (12.2) | 4,367 (12.3) |  |
| **Region** |  |  |  | <0.001 |
| South of England | 116,148 (41.4) | 100,785 (41.1) | 15,363 (43.3) |  |
| London | 27,066 (9.7) | 23,443 (9.6) | 3,623 (10.2) |  |
| Midlands and east of England | 79,274 (28.3) | 69,271 (28.3) | 10,003 (28.2) |  |
| North of England and Yorkshire | 57,974 (20.7) | 51,464 (21.0) | 6,510 (18.3) |  |
| **CCI (continuous)\*** | 1.96 (1.9) | 1.95 (1.9) | 2.03 (2.0) | <0.001 |
| **CCI (categorical)** |  |  |  | <0.001 |
| 0 | 82,406 (29.4) | 72,475 (29.6) | 9,931 (28.0) |  |
| ≥ 1 | 198,056 (70.6) | 172,488 (70.4) | 25,568 (72.0) |  |
| **Smoking status** |  |  |  | <0.001 |
| Non-smoker | 167,927 (59.9) | 147,977 (60.4) | 19,950 (56.2) |  |
| Ex-smoker | 92,507 (33.0) | 79,419 (32.4) | 13,088 (36.9) |  |
| Smoker | 20,028 (7.1) | 17,567 (7.2) | 2,461 (6.9) |  |
| **Recurrent UTI** | 71,391 (25.5) | 62,890 (25.7) | 8,501 (23.9) | <0.001 |
| **Hospital stays** |  |  |  |  |
| Discharged from hospital<br>(in prior 7 days) | 6,526 (2.3) | 5,396 (2.2) | 1,130 (3.2) | <0.001 |
| Discharged from hospital<br>(in prior 30 days) | 20,655 (7.4) | 17,682 (7.2) | 2,973 (8.4) | <0.001 |
| Number of weeks in hospital*<br>(in prior year) | 0.54 (1.9) | 0.52 (1.9) | 0.65 (2.2) | <0.001 |
| Number of admissions*<br>(in prior year) | 0.32 (0.8) | 0.31 (0.8) | 0.38 (0.9) | <0.001 |
| **Accidents & Emergencies attendances** |  |  |  |  |
| A&E attendance<br>(in prior 30 days) | 10,875 (3.9) | 8,729 (3.6) | 2,146 (6.0) | <0.001 |
| Number of attendances*<br>(in prior year) | 15,142 (5.4) | 0.44 (1.0) | 0.53 (1.2) | <0.001 |
| **Antibiotic (in prior 30 days)** | 54,077 (19.3) | 44,496 (18.2) | 9,581 (27.0) | <0.001 |
| **Index event was home visit** | 12,531 (4.5) | 9,116 (3.7) | 3,415 (9.6) | <0.001 |
| **Outcomes (within 60 days after episode start)** |  |  |  |  |
| Sepsis | 1,253 (0.4) | 1,025 (0.4) | 228 (0.6) | <0.001 |
| Hospitalisation<br>(non-sepsis, non-UTI) | 16,492 (5.9) | 13,700 (5.6) | 2,792 (7.9) | <0.001 |
| Death | 5,636 (2.0) | 4,593 (1.9) | 1,043 (2.9) | <0.001 |

Note that patients can have more than one UTI episode within the study period and will be counted separately for each of their episodes. \* Coded as a continuous variable and summarised by mean and standard deviation.

The crude odds of sepsis were higher in patients who were not treated with antibiotics immediately, compared to patients who received a prescription on the date of their first consultation for UTI (OR 1·53, 95-% CI: 1·33-1·77; Table 2). However, in the adjusted analysis we found no evidence that delaying or withholding treatment was associated with an increased likelihood of sepsis in the following 60 days (OR 1·13, 95-% CI: 0·97-1·31). The corresponding NNEH was 1,882, i.e. we would anticipate one extra case of sepsis for every 1,882 patients not treated immediately with antibiotics. The estimated lower bound of the 95% confidence interval was 830, reflecting uncertainty in the OR (upper limit not calculated). Women were less likely to develop BSI compared to men (OR 0·48, 95% CI: 0·42-0·54) and increasing age (OR 1·24, 95% CI: 1·20-1·29 per 5 years) and social deprivation (Q5 versus Q1: 1·48; 95%-CI: 1·22-1·80) were independently associated with BSI.

**Table 2:**
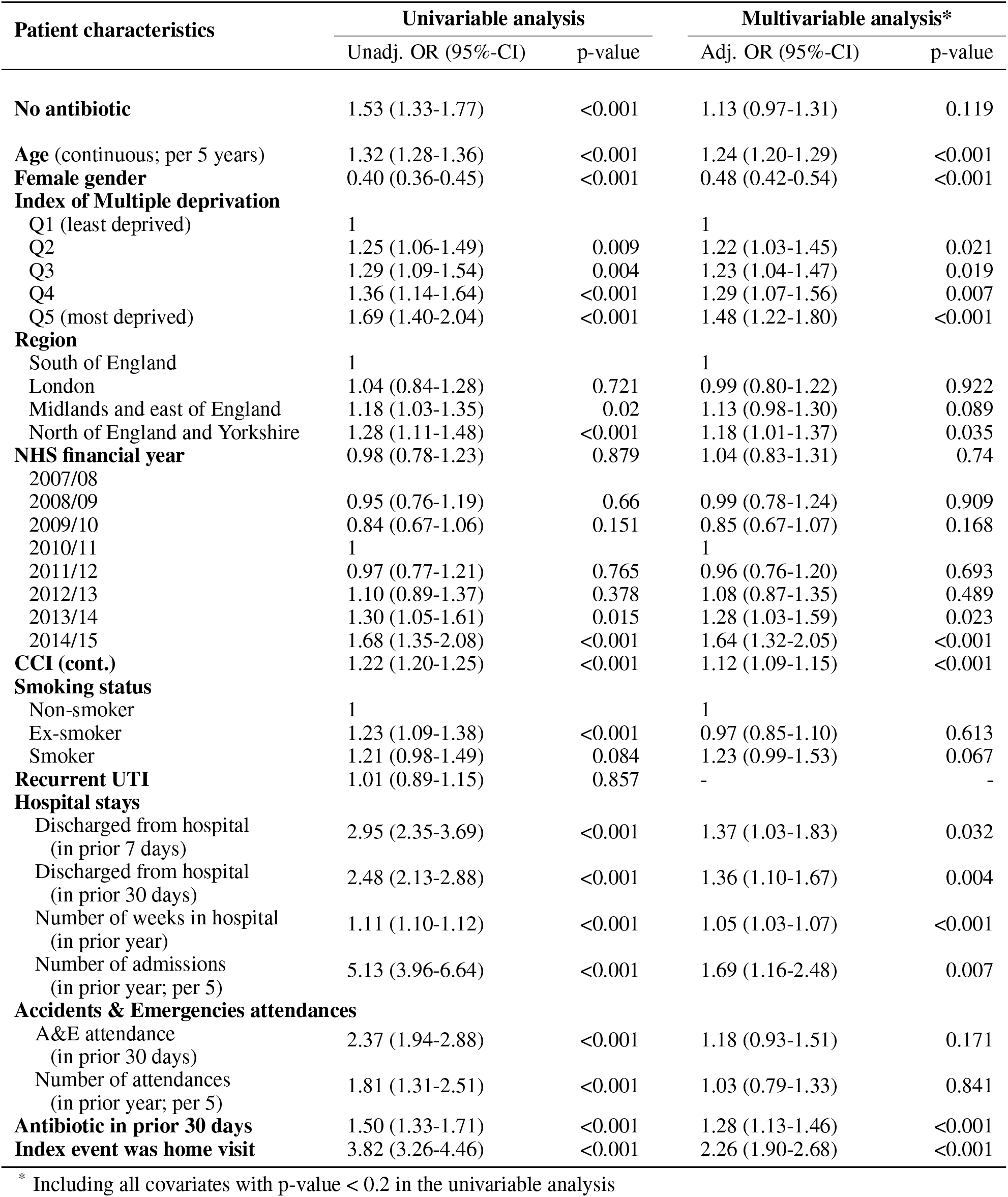
Univariable and multivariable associations between immediate antibiotic prescribing for UTI and sepsis within 60 days, adjusting for covariates using generalized estimating equations and Huber-White sandwich estimators

| Patient characteristics | Univariable analysis |  | Multivariable analysis* |  |
| --- | --- | --- | --- | --- |
|  | Unadj. OR (95%-CI) | p-value | Adj. OR (95%-CI) | p-value |
| **No antibiotic** | 1.53 (1.33-1.77) | <0.001 | 1.13 (0.97-1.31) | 0.119 |
| **Age** (continuous; per 5 years) | 1.32 (1.28-1.36) | <0.001 | 1.24 (1.20-1.29) | <0.001 |
| **Female gender** | 0.40 (0.36-0.45) | <0.001 | 0.48 (0.42-0.54) | <0.001 |
| **Index of Multiple deprivation** |  |  |  |  |
| Q1 (least deprived) | 1 |  | 1 |  |
| Q2 | 1.25 (1.06-1.49) | 0.009 | 1.22 (1.03-1.45) | 0.021 |
| Q3 | 1.29 (1.09-1.54) | 0.004 | 1.23 (1.04-1.47) | 0.019 |
| Q4 | 1.36 (1.14-1.64) | <0.001 | 1.29 (1.07-1.56) | 0.007 |
| Q5 (most deprived) | 1.69 (1.40-2.04) | <0.001 | 1.48 (1.22-1.80) | <0.001 |
| **Region** |  |  |  |  |
| South of England | 1 |  | 1 |  |
| London | 1.04 (0.84-1.28) | 0.721 | 0.99 (0.80-1.22) | 0.922 |
| Midlands and east of England | 1.18 (1.03-1.35) | 0.02 | 1.13 (0.98-1.30) | 0.089 |
| North of England and Yorkshire | 1.28 (1.11-1.48) | <0.001 | 1.18 (1.01-1.37) | 0.035 |
| **NHS financial year** | 0.98 (0.78-1.23) | 0.879 | 1.04 (0.83-1.31) | 0.74 |
| 2007/08 |  |  |  |  |
| 2008/09 | 0.95 (0.76-1.19) | 0.66 | 0.99 (0.78-1.24) | 0.909 |
| 2009/10 | 0.84 (0.67-1.06) | 0.151 | 0.85 (0.67-1.07) | 0.168 |
| 2010/11 | 1 |  | 1 |  |
| 2011/12 | 0.97 (0.77-1.21) | 0.765 | 0.96 (0.76-1.20) | 0.693 |
| 2012/13 | 1.10 (0.89-1.37) | 0.378 | 1.08 (0.87-1.35) | 0.489 |
| 2013/14 | 1.30 (1.05-1.61) | 0.015 | 1.28 (1.03-1.59) | 0.023 |
| 2014/15 | 1.68 (1.35-2.08) | <0.001 | 1.64 (1.32-2.05) | <0.001 |
| **CCI (cont.)** | 1.22 (1.20-1.25) | <0.001 | 1.12 (1.09-1.15) | <0.001 |
| **Smoking status** |  |  |  |  |
| Non-smoker | 1 |  | 1 |  |
| Ex-smoker | 1.23 (1.09-1.38) | <0.001 | 0.97 (0.85-1.10) | 0.613 |
| Smoker | 1.21 (0.98-1.49) | 0.084 | 1.23 (0.99-1.53) | 0.067 |
| **Recurrent UTI** | 1.01 (0.89-1.15) | 0.857 | - | - |
| **Hospital stays** |  |  |  |  |
| Discharged from hospital (in prior 7 days) | 2.95 (2.35-3.69) | <0.001 | 1.37 (1.03-1.83) | 0.032 |
| Discharged from hospital (in prior 30 days) | 2.48 (2.13-2.88) | <0.001 | 1.36 (1.10-1.67) | 0.004 |
| Number of weeks in hospital (in prior year) | 1.11 (1.10-1.12) | <0.001 | 1.05 (1.03-1.07) | <0.001 |
| Number of admissions (in prior year; per 5) | 5.13 (3.96-6.64) | <0.001 | 1.69 (1.16-2.48) | 0.007 |
| **Accidents & Emergencies attendances** |  |  |  |  |
| A&E attendance (in prior 30 days) | 2.37 (1.94-2.88) | <0.001 | 1.18 (0.93-1.51) | 0.171 |
| Number of attendances (in prior year; per 5) | 1.81 (1.31-2.51) | <0.001 | 1.03 (0.79-1.33) | 0.841 |
| **Antibiotic in prior 30 days** | 1.50 (1.33-1.71) | <0.001 | 1.28 (1.13-1.46) | <0.001 |
| **Index event was home visit** | 3.82 (3.26-4.46) | <0.001 | 2.26 (1.90-2.68) | <0.001 |

\* Including all covariates with p-value < 0.2 in the univariable analysis

Comorbidity, prior hospital admissions and antibiotic treatment in the prior 30 days were all associated with increased odds of sepsis. The odds of BSI were also increased in patients who received a home visit from their GP (OR 2·26, 95% CI: 1·90-2 68), including visits to care homes. We found modest evidence (p=0·047) that gender but not age modified the association between delayed or withheld antibiotics and BSI (Women: OR 1·29, 95% CI: 1·04-1 59; Men: OR 0·97, 95%CI: 0·79-1 20; Supplementary Table 2). Since we had not previously hypothesised an interaction between gender and treatment all subsequent analyses excluded interactions.

Delaying or withholding antibiotics was associated with increased mortality in the subsequent 60 days (OR 1·17, 95% CI: 1·09-1·26; Supplementary Table 3). The corresponding NNEH was 326 i.e. for every 326 (95% CI: 236-534) patients not immediately treated with antibiotics we observed one additional death within 60 days. However, in sensitivity analysis, patients who were not treated immediately with antibiotics were also more likely to have been admitted to hospital for conditions unrelated to sepsis or UTI in the 60 days following consultation (OR 1·20, 95% CI: 1·14-1·25). Restricting the analysis to each patient’s first episode of UTI supported our main findings of no association between delayed or withheld treatment and sepsis (OR 0 96, 95% CI: 0·79-1 18; Supplementary Table 4), but shortening the period of follow-up to 30 days provided some evidence of an association between delaying/withholding treatment and sepsis (OR 1·25, 95% CI: 1·06-1·48). Use of propensity scores to address residual confounding led to odds ratios for the association between delayed/withheld prescribing and sepsis that ranged from 1·10 (95%-CI: 0·94-1·27) to 1·28 (95%-CI: 1·09-1·51) depending on the method applied (Supplementary Tables 5&6).

Finally, in-depth analysis of the cause of sepsis showed that one quarter of cases had urosepsis recorded at some point during hospital admission, with urosepsis listed as the main reason for admission in just 129/1253 (10·3%) of all sepsis cases (Table 3). More than one-third of hospital-confirmed sepsis cases were attributed to non-urinary sources, mainly respiratory infections. A diagnostic code for sepsis was solely recorded in the primary care record as in 394 cases (31·4%).

**Table 3:**
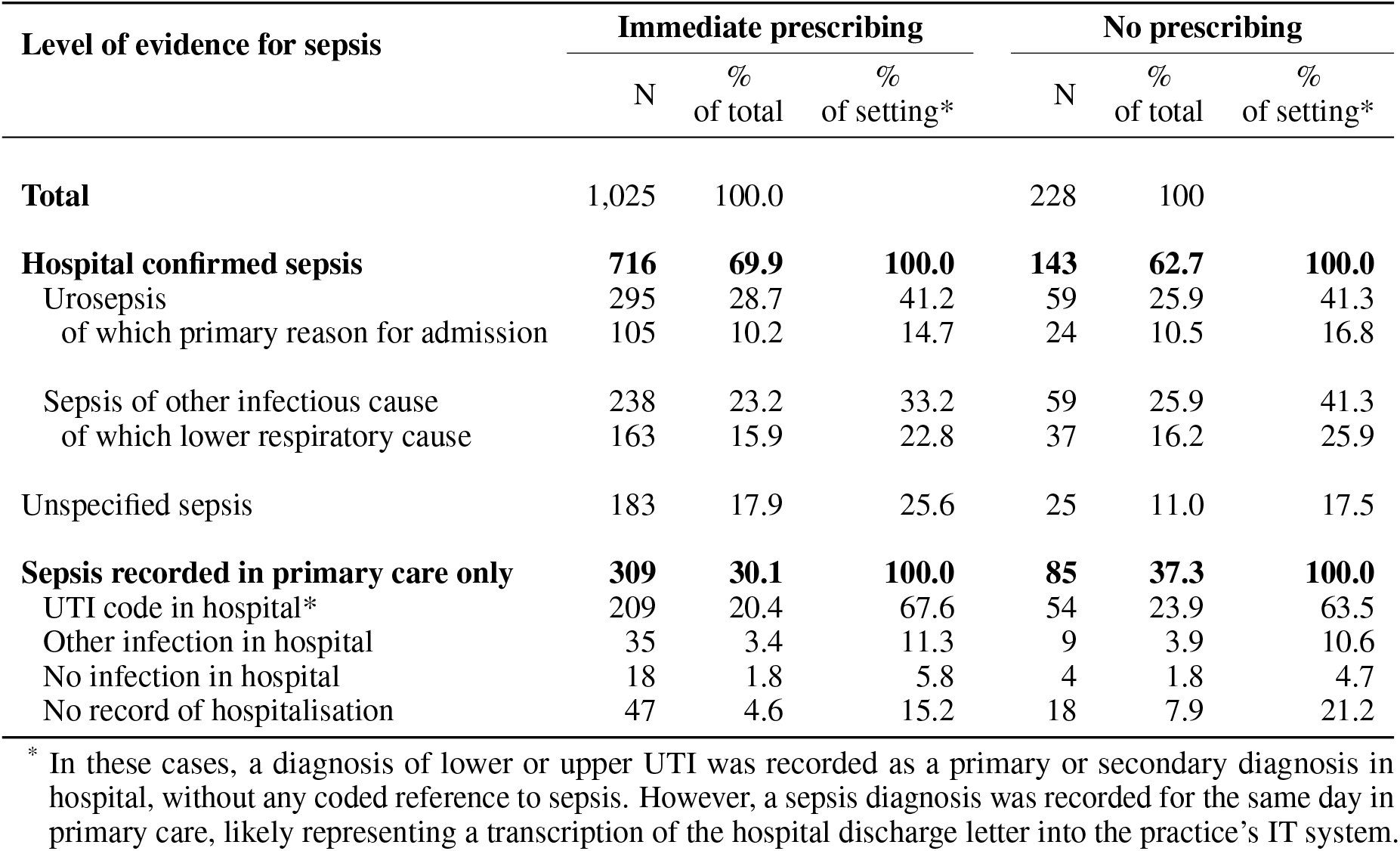
Healthcare setting and recorded cause of sepsis recorded within 60 days of episode start date

| Level of evidence for sepsis | Immediate prescribing |  |  | No prescribing |  |  |
| --- | --- | --- | --- | --- | --- | --- |
|  | N | % of total | % of setting* | N | % of total | % of setting* |
| **Total** | 1,025 | 100.0 |  | 228 | 100 |  |
| **Hospital confirmed sepsis** | **716** | **69.9** | **100.0** | **143** | **62.7** | **100.0** |
| Urosepsis | 295 | 28.7 | 41.2 | 59 | 25.9 | 41.3 |
| of which primary reason for admission | 105 | 10.2 | 14.7 | 24 | 10.5 | 16.8 |
| Sepsis of other infectious cause | 238 | 23.2 | 33.2 | 59 | 25.9 | 41.3 |
| of which lower respiratory cause | 163 | 15.9 | 22.8 | 37 | 16.2 | 25.9 |
| Unspecified sepsis | 183 | 17.9 | 25.6 | 25 | 11.0 | 17.5 |
| **Sepsis recorded in primary care only** | **309** | **30.1** | **100.0** | **85** | **37.3** | **100.0** |
| UTI code in hospital* | 209 | 20.4 | 67.6 | 54 | 23.9 | 63.5 |
| Other infection in hospital | 35 | 3.4 | 11.3 | 9 | 3.9 | 10.6 |
| No infection in hospital | 18 | 1.8 | 5.8 | 4 | 1.8 | 4.7 |
| No record of hospitalisation | 47 | 4.6 | 15.2 | 18 | 7.9 | 21.2 |

\* In these cases, a diagnosis of lower or upper UTI was recorded as a primary or secondary diagnosis in hospital, without any coded reference to sepsis. However, a sepsis diagnosis was recorded for the same day in primary care, likely representing a transcription of the hospital discharge letter into the practice's IT system.

## 4 Discussion

In this study, delaying or withholding antibiotic treatment for suspected UTI was not associated with an increased risk of BSI, but patients who did not receive antibiotics immediately were more likely to die in the following 60 days. This equates to approximately one additional death for every 300 patients aged ≥ 65 years who were not treated immediately with antibiotics.

Analysis of BSI diagnostic codes revealed that only half of sepsis cases could be linked to a UTI code, with clear evidence of a non-urinary source, such as skin or respiratory infection in more than 25% of cases. Delaying or withholding antibiotic treatment was also associated with increased hospital admissions for causes unrelated to UTI or BSI. This implies that the risk of BSI and death in some patients included in this study may have been driven by a delay in diagnosing the patient’s underlying illness, rather than a delay in initiating antibiotics for lower UTI.

### 4.1 Strengths and limitations of this study

A major strength of our analysis is the use of a large and nationally representative primary care database (>850,000 patients) linked to hospital admissions. This means our estimates can be generalised to the UK population.^14^ Linkage of the primary care dataset to HES allowed us to apply stringent criteria to identify community-onset UTI cases by differentiating new from ongoing UTI episodes and excluding cases that originated in hospital. Sensitivity analyses also support our main conclusions and highlight the limitations of diagnostic coding for sepsis.

Limitations relate to the fact that electronic health records are designed for clinical care not research. Observational studies using CPRD are at risk of confounding by indication if there are systematic differences (such as the severity of symptoms) between patients who receive a prescription and those who do not. This is particularly challenging when the exposure of interest is unevenly distributed across the study population as seen in this study (87% of patients received an immediate antibiotic versus 13% who did not). Estimates from our propensity score analysis were congruent with our main findings, but we acknowledge that residual confounding is likely.

Read codes were used to identify patients with suspected UTI, since microbiological culture of urine is usually only performed for patients with recurrent UTI or when the clinician suspects that the patients may have a drug-resistant infection. Consequently, it is likely that our cohort included patients with asymptomatic bacteriuria and/or other types of infections. Up to 40% of prescriptions for nitrofurantoin are not linked to a Read code,^18^ which suggests that we may have failed to identify some patients who were treated immediately with antibiotics. This also highlights challenges associated with using Read codes to infer the date of infection onset.

Cases of sepsis were identified from ICD-10 code or Read codes and we found that almost one-third of sepsis diagnoses were only recorded in primary care. It is difficult to disentangle the reasons for this since almost all cases of sepsis are managed in hospital. Patients may have received treatment for sepsis abroad or in a non-NHS setting, or information from the discharge letter may have been used to infer the diagnosis of sepsis. Linkage of microbiological data to HES/CPRD would enable more accurate estimation of the proportion of sepsis cases that could be attributed to a urinary source.

### 4.2 Comparison with existing literature

Previous studies of alternatives to antibiotics or delayed prescribing for community-onset UTI have usually focused on women aged 18-70 years. A systematic review of trials in young non-pregnant women reported that antibiotic treatment was associated with more rapid resolution of urinary symptoms and microbiological cure based on urine culture, compared to placebo,^19^ but not with reduced incidence of pyelonephritis. Delayed prescribing has also been safely used in low-risk women with uncomplicated UTI,^20^ provided there is adequate safety-netting and self-care advice.^2^ For example, in a trial comparing treatments for uncomplicated UTI in women aged 18-65 years,^21^ women receiving ibuprofen had a higher burden of symptoms but considerably less antibiotic exposure compared to women treated with fosfomycin (incident risk reduction 66·5%, 95%-CI: 58·8-74·4%; P<0·001), and two-thirds of patients in the ibuprofen group recovered without antibiotics.

Whilst the efficacy and safety of delayed prescribing for respiratory tract infections in primary care is well-established,^22^ implementing similar approaches for UTI is controversial due to concerns around prolongation of symptoms, and the potential risk of antimicrobial resistance and complicated UTI resulting from inadequate antibiotic therapy. These issues are particularly relevant in elderly patients who have the highest incidence of community-onset UTI, but also the highest incidence of *E. coli* BSI,^3^ which may be a consequence of suboptimal antibiotic treatment in primary care.

With the exception of Gharbi *et al*., few studies have evaluated the use of delayed prescribing or alternatives to antibiotics in older adults. These patients arguably have the most to gain from prudent antibiotic prescribing, due to their increased risk of adverse outcomes related to antibiotic use^23^ and high prevalence of asymptomatic bacteriuria.^6^ The major barrier to delaying or withholding antibiotics in these individuals is the risk of UTI-related complications, as reported by Gharbi *et al*.^12^ Our analysis, and concerns raised by other research groups, call these findings into question. We find no evidence of an association between delaying or withholding antibiotics and sepsis, but some evidence of increased mortality. The discrepancy between our analysis and that conducted by Gharbi *et al*. is likely to relate to the different approaches used to define community-onset UTI, residual confounding and the limitations of coding in electronic health records.

### 4.3 Clinical, policy and research implications

This population-based study highlights uncertainty around the safety of delaying or withholding antibiotic treatment for suspected UTI in patients aged ≥ 65 years. This lack of clarity may make GPs less willing to consider the use of delayed prescribing in older adults with suspected UTI, increasing the likelihood that these patients will be exposed to antibiotics unnecessarily. For researchers, our findings highlight methodological challenges associated with defining the onset of infection using electronic health records and the need for linkage of microbiological datasets to HES/CPRD.

### 4.4 Conclusion

The safety of delaying or withholding antibiotics in adults aged ≥ 65 years with suspected UTI is uncertain. Adverse consequences of antibiotic treatment in this population and the public health need to tackle antibiotic resistance advocate strongly for a trial to address this uncertainty.

## Data Availability

Clinical Practice Research Datalink (CPRD), Hospital Episode Statistics (HES) and Office for National Statistics (ONS) data cannot be directly shared by the researchers but are available directly from CPRD and NHS Digital subject to standard conditions. All statistical code is available from https://github.com/prockenschaub/CPRD_UTI_sepsis_elderly.

## Acknowledgements

The authors are grateful to Dr Peter Dutey-Magni for helpful comments during the design and execution of the analysis. This work was supported by the Economic and Social Research Council (ES/P008321/1). Laura Shallcross was funded by a National Institute of Health Research (NIHR) Clinician Scientist award (CS-2016-007) for this research project. Andrew Hayward was funded by an NIHR Senior Investigator award for this project. This publication presents independent research funded by the NIHR. The views expressed are those of the authors alone and not necessarily those of the NHS, the NIHR or the Department of Health and Social Care. Neither the funders nor the study sponsor played any role in the design, analysis or reporting of the study. This study was carried out as part of the CALIBER^*©*^ programme (https://www.ucl.ac.uk/health-informatics/caliber). CALIBER, led from the UCL Institute of Health Informatics, is a research resource consisting of anonymised, coded variables extracted from linked electronic health records, methods and tools, specialised infrastructure, and training and support. This study is based in part on data from the Clinical Practice Research Datalink obtained under licence from the UK Medicines and Healthcare products Regulatory Agency. The data is provided by patients and collected by the NHS as part of their care and support. This study is further based on data from the Hospital Episode Statistics. Copyright^*©*^ (2019), re-used with the permission of The Health & Social Care Information Centre. All rights reserved.

## Contributors

LS, PR, NF, IH and AH designed the study. PR performed all statistical analyses reported in this study, which was independently replicated by RB. LS and PR interpreted the data and wrote the draft manuscript. All authors revised, edited and approved the final manuscript.

